# Sex-specific associations between metabolic dysregulation and knee pain: a 9-year population-based cohort study

**DOI:** 10.64898/2026.05.21.26353831

**Authors:** Ivan Shirinsky, Alexandra Makogon, Elena Shakhtshneider, Diana Denisova, Elena Belyaevskaya, Valery Shirinsky

## Abstract

**Introduction:** Knee pain is a highly prevalent condition in the general population and is more common than knee osteoarthritis. Population-based evidence linking metabolic dysfunction to knee pain remains limited, and data on sex-specific effects are scarce. Therefore, we examined sex-specific associations between metabolic dysregulation and knee pain in a population-based cohort.

**Method:** We analyzed data from a population-based cohort of 1,512 adults (mean age 37.2 years at baseline), of whom 250 completed follow-up after a mean of 9.4 years. Metabolic dysfunction was assessed using a continuous MetS severity score (cMetS) derived from waist circumference, triglycerides, HDL cholesterol, fasting glucose, and systolic blood pressure. Knee pain at follow-up was defined using a combined measure based on a standardized question and a body manikin. Logistic regression models were used to examine associations between baseline cMetS and knee pain, including interaction analyses by sex.

**Results:** At follow-up, 28.5% of participants reported knee pain. Higher baseline cMetS was associated with increased odds of knee pain in males (odds ratio [OR] 1.41, 95% confidence interval [CI] 1.17–1.69) but not in females (OR 0.94, 95% CI 0.84–1.07), with evidence of interaction by sex (interaction P < 0.001). Findings were consistent across sensitivity analyses.

**Conclusions:** These results indicate that metabolic dysfunction is associated with knee pain in males but not in females, suggesting sex-specific mechanisms linking metabolic dysfunction and knee pain.

**Key-points:**

- Population-based cohort study with approximately 9 years of follow-up
- Metabolic dysfunction was assessed using a continuous metabolic syndrome severity score (cMetS)
- Higher baseline cMetS predicted knee pain in males but not in females
- Significant interaction by sex suggests sex-specific mechanisms linking metabolic dysfunction and knee pain

## Introduction

Knee pain is one of the most common musculoskeletal complaints in adults [1]. Although it is often considered a symptom of osteoarthritis (OA), population-based data suggest that knee pain is substantially more prevalent than OA. For example, in the UK Biobank, the number of participants reporting knee pain was approximately twice as high as those with OA [2]. This discrepancy suggests that knee pain may represent an important musculoskeletal outcome that cannot be fully explained by structural joint disease alone [3]. Despite its high prevalence and clinical importance, comparatively fewer population-based studies have examined knee pain as a primary outcome [4], with most studies focusing on OA [5].

Metabolic syndrome (MetS) is characterized by a clustering of abdominal obesity, dyslipidaemia, hyperglycaemia, and hypertension. It is well established as a risk factor for cardiovascular disease and type 2 diabetes [6]. In addition, MetS has been linked to low-grade systemic inflammation, adipokine dysregulation, and metabolic alterations that may adversely affect joint tissues and pain signaling pathways [7]. Given these mechanisms, MetS may plausibly contribute to musculoskeletal outcomes, including knee pain.

However, the relationship between MetS and knee pain remains incompletely understood. A meta-analysis questioned the association between MetS and knee OA [8], whereas more recent longitudinal evidence from a population-based cohort suggested an association between MetS and knee pain [9].

Metabolic risk factors may exert differential effects across population subgroups. Many cardiometabolic traits vary substantially between males and females [10]. Pain processing and the transition to chronic pain also differ between males and females, reflecting distinct underlying mechanisms [11]. Recent methodological recommendations further emphasize the importance of considering sex as a biological variable in pain research [12].

To date, population-based evidence on sex-specific associations between metabolic factors and knee pain remains limited. Much of the available evidence comes from the Rotterdam Study, in which MetS and several of its components were associated with incident chronic knee pain in males but not in females, suggesting potential sex-specific effects. However, these data were derived from an older population, and it remains unclear whether similar patterns are present in younger adults or are consistent across populations [13].

Traditional definitions of MetS are based on binary criteria, which may limit their ability to capture the full spectrum of metabolic risk. To address these limitations, continuous MetS severity scores have been proposed, including those based on standardized component z-scores. These scores allow for a more nuanced assessment of metabolic burden [14].

To our knowledge, no studies have examined the relationship between a continuous metabolic syndrome score and knee pain in a population-based setting. Our objective was to examine this association in a population-based adult cohort, with a particular focus on potential sex differences.

## Method

### Study design and population

This study used data from a population-based adult cohort established in one administrative district of Novosibirsk, Russia. The parent study was originally designed to investigate cardiovascular and metabolic risk factors in the general population. Participants were recruited between 2013 and 2017 using a standardized epidemiological protocol, as described previously [15]. A total of 1,512 individuals agreed to participate and were included in the baseline cohort. The primary aim of the parent study was to assess cardiovascular and metabolic risk factors in adults from the general population.

At baseline, all participants underwent standardized assessments, including anthropometric measurements and biochemical analyses. Body mass index (BMI) was calculated as weight (kg) divided by height squared (m²). Sex was treated as a biological variable (male/female) based on data recorded at baseline. Gender identity was not assessed in this study. Fasting blood samples were collected after an overnight fast of at least 12 hours. Serum levels of glucose, total cholesterol, triglycerides, and high-density lipoprotein cholesterol were measured using standard enzymatic methods. Smoking status, education level, physical activity, and employment status were assessed by questionnaire. Self-rated health was assessed using a single-item 5-category scale (very good, good, fair, poor, very poor).

A follow-up examination of the cohort was initiated approximately 10 years after baseline (from 2023 onwards). As part of this follow-up, an extended assessment protocol focusing on musculoskeletal health, knee pain, and OA was implemented as a dedicated sub-study within the cohort framework. Follow-up assessments were initiated in March 2023 and are ongoing. The present analysis represents the cohort at the time of data extraction (March 2026) and includes participants who had completed the follow-up assessment by that time. The mean follow-up duration was 9.4 ± 1.2 years. The analytical protocol for the musculoskeletal follow-up study is publicly available on the Open Science Framework (OSF) (https://osf.io/zfeqm/) [16]. The available version provides a summary of the study design and planned analyses.

### Exposure

The primary exposure was a continuous metabolic syndrome severity score (cMetS), constructed at baseline as a composite measure of metabolic risk. The score included waist circumference, triglycerides, high-density lipoprotein cholesterol (HDL-C), fasting glucose, and systolic blood pressure. To account for age- and sex-related differences, each component was regressed on age and sex, and standardized residuals (z-scores) were obtained. These residuals represent the deviation of each individual’s observed value from that expected based on age and sex. The overall cMetS score was calculated as the sum of standardized residuals of the individual components, with HDL-C included inversely (multiplied by −1), so that higher values reflected a less favorable metabolic profile, consistent with previously described methods [17]. The exposure was defined at baseline for longitudinal analyses and at visit 2 for cross-sectional analyses.

### Outcome

Assessment of musculoskeletal pain was performed using a body manikin, on which participants were asked to indicate areas of pain experienced for at least one week during the previous month [18].

Knee pain was assessed using a standardized question: “Have you had pain in or around the knee on most days during the last month?” Participants answering “yes” were classified as having knee pain [19].

To improve sensitivity, information from the body manikin and the direct question on knee pain was combined. Participants were considered to have knee pain (broad definition) if they reported pain in the knee region on the manikin and/or answered positively to the direct question.

### Statistical analysis

Descriptive statistics were used to summarize participant characteristics at baseline. Continuous variables are presented as mean ± standard deviation (SD), and categorical variables as counts and percentages.

The outcome of interest was knee pain at follow-up. Associations between metabolic exposures and knee pain were examined using logistic regression models, with results presented as odds ratios (ORs) and 95% confidence intervals (CIs).

The primary analyses were longitudinal. Secondary cross-sectional analyses examined associations between visit 2 cMetS and knee pain at visit 2. For both longitudinal and cross-sectional analyses, two models were fitted: a base model including cMetS, sex, and age, and a second model including a cMetS × sex interaction term to assess effect modification. Separate logistic regression models were also fitted for each individual baseline metabolic component, entered separately as age- and sex-standardized residual z-scores. These models were adjusted for age and sex.

To evaluate potential selection bias due to differential participation in follow-up at the time of analysis, baseline characteristics of participants who had completed the follow-up examination were compared with those who had not completed the follow-up assessment at the time of analysis using standardized mean differences (SMDs), with values >0.1 indicating meaningful imbalance.

### Sensitivity analyses

To examine the robustness of the longitudinal findings to potential selection bias due to differential participation in the follow-up assessment at the time of analysis, inverse probability weighting (IPW) was applied in sensitivity analyses [20]. Response probabilities were estimated in the full baseline sample using logistic regression with participation in the follow-up visit as the dependent variable. Two response models were used. One model included baseline cardiometabolic variables together with available sociodemographic and clinical predictors of follow-up participation. The second model included baseline age, sex, waist circumference, triglycerides, HDL cholesterol, fasting glucose, and systolic blood pressure. Inverse probability weights were then applied to the primary logistic regression model including cMetS, sex, age, and the cMetS × sex interaction term. Results were compared with the main unweighted analysis. As an additional sensitivity analysis, inverse probability weights were truncated at the 1st and 99th percentiles to limit the influence of extreme weights [21].

As a further sensitivity analysis, the main longitudinal model was additionally adjusted for baseline body mass index (BMI). Because waist circumference was included as a component of the cMetS score, BMI adjustment was treated as a sensitivity analysis due to potential overadjustment and collinearity.

Missing data were handled using complete-case analysis within each model. Statistical significance was defined as a two-sided p value <0.05. All analyses were performed in R [22]. ***Sample size***

The study was designed as a prospective observational cohort, and the sample size was determined pragmatically based on recruitment feasibility rather than a predefined statistical calculation.

### Ethics

The study was approved by the local ethics committee, and all participants provided informed consent.

### Reporting

All analyses were conducted in accordance with established methodological principles for observational studies and are reported following the STROBE Statement [23]. A completed STROBE checklist is provided in the Supplementary Material.

### Patient and public involvement

Patients and members of the public were not involved in the design, conduct, reporting, or dissemination planning of this research.

## Results

Of 1,512 participants at baseline, 250 completed follow-up assessments at the time of analysis. Knee pain status at follow-up was available for 249 participants, of whom 71 (28.5%) reported knee pain. Among these, 30 participants reported pain on the manikin only, 5 on the direct question only, and 36 on both. Baseline characteristics according to knee pain status are presented in Table 1.

**Table 1.** Baseline characteristics of participants according to knee pain status at 9-year follow-up.

| **Variable** | **Overall** | **No knee pain** | **Knee pain** |
| --- | --- | --- | --- |
| Participants, n | 249 | 178 | 71 |
| Age, years | 37.2 ± 5.9 | 37.0 ± 5.8 | 37.7 ± 6.2 |
| Female, n (%) | 129 (51.8%) | 84 (47.2%) | 45 (63.4%) |
| BMI, kg/m<sup>2</sup> | 26.0 ± 4.7 | 25.7 ± 4.7 | 26.8 ± 4.7 |
| Waist circumference, cm | 86.7 ± 12.8 | 86.0 ± 12.5 | 88.4 ± 13.4 |
| Triglycerides, mmol/L | 1.26 ± 0.98 | 1.21 ± 0.79 | 1.39 ± 1.32 |
| HDL-C, mmol/L | 1.33 ± 0.32 | 1.32 ± 0.31 | 1.35 ± 0.35 |
| Glucose, mmol/L | 5.55 ± 0.58 | 5.55 ± 0.58 | 5.56 ± 0.58 |
| Systolic BP, mmHg | 119.9 ± 14.1 | 120.4 ± 13.9 | 118.6 ± 14.7 |
| MetS by IDF, n (%) | 69 (27.7%) | 46 (25.8%) | 23 (32.4%) |
| cMetS | -0.15 ± 3.04 | -0.35 ± 2.80 | 0.35 ± 3.53 |

Note: Continuous variables are presented as mean ± standard deviation and categorical variables as number (%). Knee pain status was missing for one participant at follow-up; this individual was excluded from the analysis. MetS by IDF was defined using clinical and laboratory criteria; information on antihypertensive treatment was not included. Abbreviations: BMI = body mass index; HDL-C = high-density lipoprotein cholesterol; BP = blood pressure; MetS = metabolic syndrome; cMetS = continuous metabolic syndrome risk score; IDF = International Diabetes Federation.

Participants who reported knee pain at follow-up were more frequently female (63.4% vs 47.2% in those without knee pain), whereas mean age was similar between groups. Individuals with knee pain tended to have slightly higher body mass index (26.8 vs 25.7 kg/m²) and waist circumference (88.4 vs 86.0 cm) at baseline. Baseline triglyceride levels were modestly higher in participants with knee pain (1.39 vs 1.21 mmol/L), whereas HDL-C and glucose levels were comparable between groups, and systolic blood pressure was slightly lower. The prevalence of MetS according to the International Diabetes Federation criteria was higher among participants with knee pain (32.4% vs 25.8%), and cMetS was also higher (0.35 vs −0.35).

In models without a cMetS × sex interaction term, no clear overall association between baseline cMetS and knee pain at follow-up was observed (Supplementary Table S1). Sex-specific estimates from the model including interaction term are presented in Table 2. Among males, higher baseline cMetS was associated with increased odds of knee pain at follow-up (OR per 1-unit increase 1.41, 95% CI 1.17 to 1.69), whereas no association was observed among females (OR 0.94, 95% CI 0.84 to 1.07). There was evidence of interaction by sex (P < 0.001), indicating that the association between cMetS and knee pain differed between males and females.

**Table 2.**
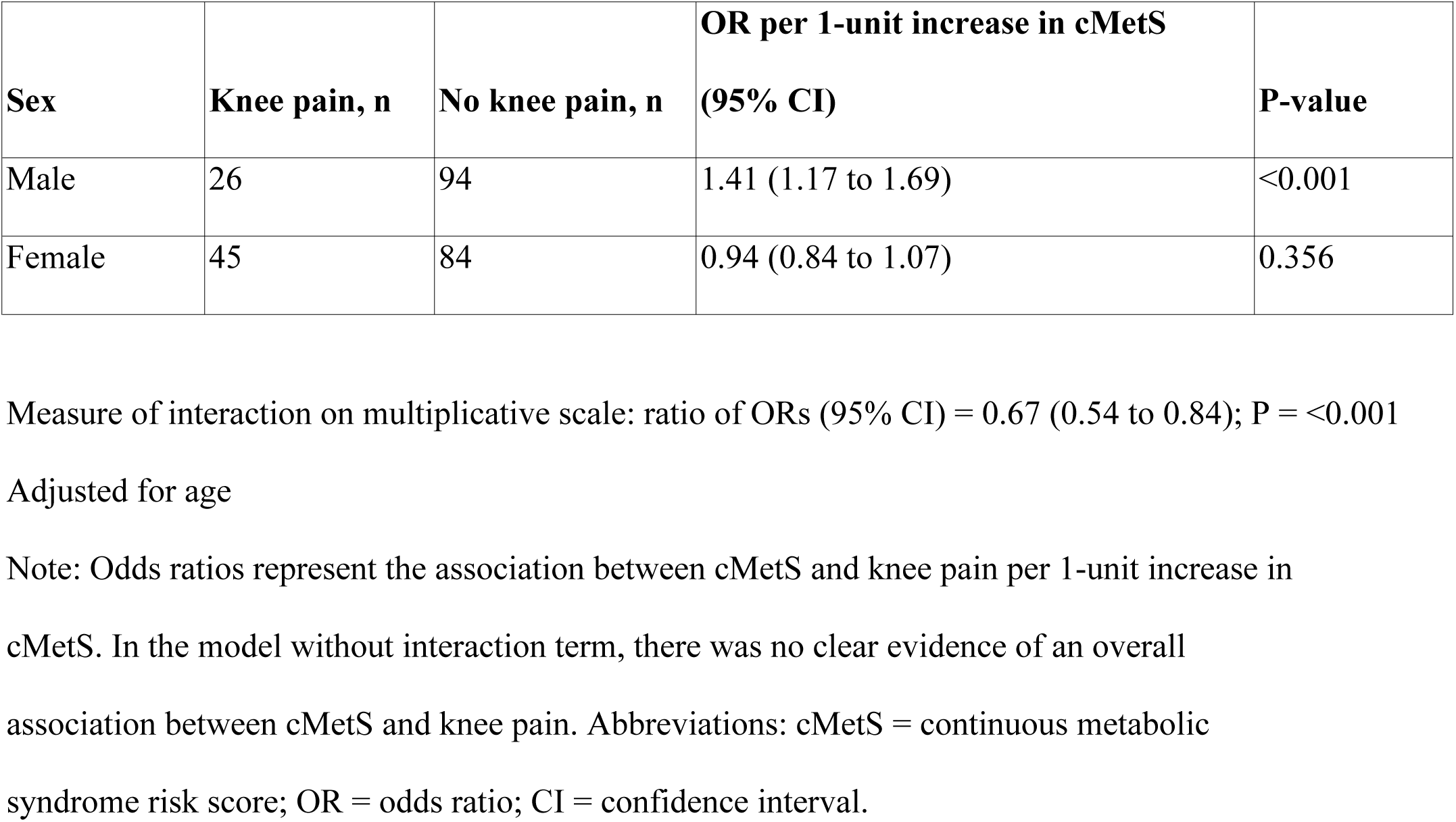
Sex-specific association between baseline cMetS and knee pain at 9-year follow-up.

Sex-specific associations between individual baseline components of cMetS and knee pain at follow-up are presented in Supplementary Table S2. Among males, several components were associated with increased odds of knee pain, including waist circumference, glucose, and systolic blood pressure, whereas higher HDL-C was associated with lower odds. In contrast, no consistent pattern of associations was observed among females, although there was evidence of interaction by sex for multiple components.

We next examined cross-sectional associations between overall metabolic dysregulation, as captured by cMetS, and knee pain at follow-up (Table 3). Higher cMetS was associated with increased odds of knee pain in males (OR 1.25 per 1-unit increase, 95% CI 1.08 to 1.46), but not in females (OR 0.94, 95% CI 0.83 to 1.07). Consistent with longitudinal analyses, there was evidence of interaction by sex (P < 0.001).

**Table 3.** Sex-specific cross-sectional association between cMetS and knee pain at 9-year follow-up.

| Sex | Knee pain, n | No knee pain, n | OR per 1-unit increase in cMetS (95% CI) | P-value |
| --- | --- | --- | --- | --- |
| Male | 26 | 94 | 1.25 (1.08 to 1.46) | 0.004 |
| Female | 45 | 84 | 0.94 (0.83 to 1.07) | 0.345 |

Measure of interaction on multiplicative scale: ratio of ORs (95% CI) = 0.75 (0.62 to 0.92); P = 0.005
Adjusted for age
Note: Odds ratios represent the cross-sectional association between cMetS and knee pain per 1-unit increase in cMetS at 9-year follow-up. Abbreviations: cMetS = continuous metabolic syndrome score; OR = odds ratio; CI = confidence interval.

To assess potential selection bias, baseline characteristics were compared between participants who completed the follow-up assessment and those not assessed at follow-up (Supplementary Table S3). Overall, baseline characteristics were broadly similar between groups, with SMDs below 0.2 for most variables, suggesting only small differences. There was a tendency for participants who completed follow-up to have slightly higher educational attainment, be more frequently employed, and report better self-rated health and higher levels of physical activity, whereas current smoking was more common among those not assessed at follow-up.

### Sensitivity analyses

To further assess potential selection bias, predictors of follow-up participation were examined and inverse probability weights (IPW) were derived (Supplementary Table S4). Older age and higher educational attainment were associated with a higher likelihood of follow-up participation, whereas lower physical activity was associated with reduced participation. Most cardiometabolic variables were not strongly associated with follow-up, and the distribution of IPW did not indicate substantial instability.

Sensitivity analyses using IPW yielded results consistent with the main analysis (Supplementary Table S5), with a persistent positive association between baseline cMetS and knee pain in males and a slight inverse association in females. Evidence of interaction by sex was consistent across all models. Analyses using truncated IPW (1st and 99th percentiles) produced similar results. Additional sensitivity analyses adjusting for BMI also yielded consistent findings (Supplementary Table S6).

## Discussion

In this population-based cohort study with a mean follow-up of 9.4 years, we observed sex-specific differences in the association between metabolic dysregulation and knee pain. Higher baseline metabolic dysregulation, as assessed by a cMetS, was associated with increased odds of knee pain among males, whereas no consistent association was observed among females. These findings were supported by evidence of interaction by sex and were consistent across cross-sectional and longitudinal analyses.

Analyses of individual components of metabolic dysregulation showed a similar pattern, with several cardiometabolic factors associated with knee pain in male but not in females.

Importantly, the main findings were robust in multiple sensitivity analyses, including inverse probability weighting to account for potential selection bias and models adjusted for body mass index. Our findings suggest potential sex-specific mechanisms linking metabolic dysfunction and knee pain.

Metabolic changes are associated with various types of chronic pain [24]. The underlying mechanisms may include low-grade systemic inflammation, adipokine imbalance, and metabolic alterations affecting joint tissues and nociceptive signaling. Such processes may increase peripheral nociceptive drive and contribute to pain generation [25].

Several studies have examined sex-specific effects of metabolic dysfunction on pain or OA in different locations including low back, knee, hand, as well as diffuse pain. Overall, the findings are heterogeneous and suggest that sex-specific effects may vary depending on the pain phenotype and anatomical location. Thus, in a large Japanese cross-sectional study, MetS was associated with low back pain in females but not men [26]. For knee and hand OA, the pattern was mixed and BMI-dependent. In the Rotterdam cohort, MetS was associated with an increased risk of incident chronic knee pain in men, and most associations weakened after BMI adjustment [13]. In contrast, a females-only Chingford study found that MetS was associated with painful interphalangeal hand OA after age and BMI adjustment, while the knee association disappeared once BMI was included [27]. For more diffuse pain, females again appeared more metabolically vulnerable. A three-year Korean cohort found that total fat mass and the fat:muscle ratio were associated with new or persistent pain among females only [28]. Taken together, these findings suggest that the relationship between metabolic dysfunction and pain is context-dependent and may reflect differences in the relative contribution of peripheral and central mechanisms.

Our findings are in line with those from the Rotterdam Study [13] by demonstrating a similar sex-specific pattern. However, our study population was substantially younger (mean age approximately 37 years vs 63 years in the Rotterdam Study), suggesting that metabolic influences on knee pain may be present earlier in the disease course. In addition, the use of a continuous measure of metabolic dysfunction may have increased sensitivity to detect associations.

Our results highlight the importance of considering sex as a biological variable in studies of pain and metabolic risk. Sex differences in the association between metabolic factors and knee pain may reflect differences in underlying pain mechanisms [29,30]. Sex differences in pain processing and chronic pain susceptibility may contribute to the observed associations [31]. In females, a greater contribution of central pain mechanisms may attenuate the relative contribution of metabolic factors. These central mechanisms may include enhanced central sensitization and altered descending pain modulation. In contrast, in men, where baseline pain sensitivity may be lower, pain mechanisms may be more influenced by metabolic and inflammatory processes [32]. Such differences are consistent with a framework in which metabolic factors contribute more strongly to pain phenotypes characterized by greater peripheral input.

These findings have implications for mechanistic stratification of pain rather than direct clinical decision-making. Future studies incorporating quantitative sensory testing or other measures of central pain processing may help clarify how metabolic factors interact with central mechanisms in shaping pain outcomes.

Our study has several limitations. First, no formal a priori sample size calculation was performed, and the number of participants with available follow-up data was relatively small, which may have limited statistical power. Follow-up assessments are ongoing, and the present analysis was based on participants who had completed the follow-up visit at the time of analysis. As a result, non-participation reflects temporary or partial follow-up rather than definitive loss to follow-up. To address potential selection bias arising from this, we applied inverse probability weighting, and results were consistent across analyses. Third, knee pain was assessed only at follow-up, precluding evaluation of temporal changes, limiting causal interpretation, and preventing distinction between incident and persistent knee pain. Fourth, the composite outcome combined two items with different pain duration thresholds (≥1 week vs most of the past month), potentially capturing overlapping but not identical aspects of knee pain, which may have introduced some heterogeneity in the case definition. Finally, data on race/ethnicity were not collected, which may limit generalizability to other populations. Residual confounding by lifestyle or occupational factors may also be present.

In conclusion, metabolic dysfunction was associated with knee pain in males but not in females, suggesting sex-specific differences in underlying pain mechanisms. Further studies are needed to clarify how metabolic and central pain mechanisms interact in shaping pain outcomes.

## Supporting information

STROBE checklist

## Statements and Declarations

### Data availability

Individual-level data are not publicly available due to institutional regulations governing access to participant data.

### Code availability

The analytical code and materials required to reproduce the reported analyses are available at GitHub (https://github.com/ivan-shirinsky/mets-knee-pain-analysis) and archived at Zenodo (https://doi.org/10.5281/zenodo.19688520).

### CRediT authorship contribution statement

Ivan V. Shirinsky: Conceptualization; Methodology; Formal analysis; Software; Data curation; Writing – original draft; Writing – review & editing; Project administration; Supervision.

Alexandra Makogon: Investigation; Data curation; Writing – review & editing; Validation. Elena Shakhtshneider: Investigation; Data curation; Writing – review & editing; Validation. Diana Denisova: Conceptualization; Methodology; Resources; Project administration; Writing – review & editing; Validation.

Elena Belyaevskaya: Investigation; Data curation; Writing – review & editing; Validation. Valery V. Shirinsky: Conceptualization; Methodology; Writing – review & editing; Supervision. All authors critically revised the manuscript, approved the final version, and agree to be accountable for all aspects of the work.

### Declaration of generative AI and AI-assisted technologies in the manuscript preparation process

During the preparation of this work, the authors used ChatGPT (OpenAI) for language editing and limited programming support. All analyses, code, interpretations, and conclusions were reviewed and verified by the authors.

## References

1. Bunt CW, Jonas CE, Chang JG. Knee Pain in Adults and Adolescents: The Initial Evaluation. American family physician. 2018;98:576–85.

2. Zengini E, Hatzikotoulas K, Tachmazidou I, Steinberg J, Hartwig FP, Southam L, et al. Genome-wide analyses using UK Biobank data provide insights into the genetic architecture of osteoarthritis. Nature genetics. 2018;50:549–58. 10.1038/s41588-018-0079-y

3. Meng W, Adams MJ, Palmer CNA, The 23andMe Research Team, Agee M, Alipanahi B, et al. Genome-wide association study of knee pain identifies associations with GDF5 and COL27A1 in UK Biobank. Communications biology. 2019;2:321. 10.1038/s42003-019-0568-2

4. Collier TS, Hughes T, Chester R, Callaghan MJ, Selfe J. Prognostic factors associated with changes in knee pain outcomes, identified from initial primary care consultation data. A systematic literature review. Annals of medicine. 2023;55:401–18. 10.1080/07853890.2023.2165706

5. Neogi T. The epidemiology and impact of pain in osteoarthritis. Osteoarthritis and cartilage. 2013;21:1145–53. 10.1016/j.joca.2013.03.018

6. Saklayen MG. The Global Epidemic of the Metabolic Syndrome. Current hypertension reports. 2018;20:12. 10.1007/s11906-018-0812-z

7. Berenbaum F. Osteoarthritis as an inflammatory disease (osteoarthritis is not osteoarthrosis!). Osteoarthritis and cartilage. 2013;21:16–21. 10.1016/j.joca.2012.11.012

8. Nie D, Yan G, Zhou W, Wang Z, Yu G, Liu D, et al. Metabolic syndrome and the incidence of knee osteoarthritis: A meta-analysis of prospective cohort studies. Wang Y, editor. Plos one. 2020;15:e0243576. 10.1371/journal.pone.0243576

9. Singh A, Fraser B, Venn A, Blizzard L, Jones G, Ding C, et al. Trajectory of metabolic syndrome and its association with knee pain in middle-aged adults. Diabetes & metabolic syndrome: Clinical research & reviews. 2023;17:102916. 10.1016/j.dsx.2023.102916

10. Zhernakova DV, Sinha T, Andreu-Sánchez S, Prins JR, Kurilshikov A, Balder J-W, et al. Age-dependent sex differences in cardiometabolic risk factors. Nature cardiovascular research. 2022;1:844–54. 10.1038/s44161-022-00131-8

11. Mogil JS. Qualitative sex differences in pain processing: Emerging evidence of a biased literature. Nature reviews neuroscience. 2020;21:353–65. 10.1038/s41583-020-0310-6

12. Finn DP, McGuire BE, Beggs S, Boerner KE, Davis KD, Defrin R, et al. Recommendations for the inclusion and study of sex and gender in research. Nature neuroscience. 2026;29:256–66. 10.1038/s41593-025-02164-1

13. Szilagyi I, Nguyen N, Boer C, Schiphof D, Ahmadizar F, Kavousi M, et al. Metabolic syndrome, radiographic osteoarthritis progression and chronic pain of the knee among men and women from the general population: The Rotterdam study. Seminars in arthritis and rheumatism. 2024;69:152544. 10.1016/j.semarthrit.2024.152544

14. DeBoer MD, Gurka MJ. Clinical utility of metabolic syndrome severity scores: Considerations for practitioners. Diabetes, metabolic syndrome and obesity: Targets and therapy. 2017;Volume 10:65–72. 10.2147/DMSO.S101624

15. Ragino YI, Oblaukhova VI, Denisova DV, Kovalkova NA. Abdominal obesity and other components of metabolic syndrome among the young population of Novosibirsk. The siberian medical journal. 2020;35:167–76. 10.29001/2073-8552-2020-35-1-167-176

16. Shirinsky I, Shirinsky V. Study protocol for the musculoskeletal follow-up of a population-based cohort in Novosibirsk. Open Science Framework;

17. Okosun IS, Lyn R, Davis-Smith M, Eriksen M, Seale P. Validity of a Continuous Metabolic Risk Score as an Index for Modeling Metabolic Syndrome in Adolescents. Annals of epidemiology. 2010;20:843–51. 10.1016/j.annepidem.2010.08.001

18. Hunt I. The prevalence and associated features of chronic widespread pain in the community using the ‘Manchester’ definition of chronic widespread pain. Rheumatology. 1999;38:275–9. 10.1093/rheumatology/38.3.275

19. Fernandes GS, Sarmanova A, Warner S, Harvey H, Akin-Akinyosoye K, Richardson H, et al. Knee pain and related health in the community study (KPIC): A cohort study protocol. Bmc musculoskeletal disorders. 2017;18:404. 10.1186/s12891-017-1761-4

20. Doidge JC. Responsiveness-informed multiple imputation and inverse probability-weighting in cohort studies with missing data that are non-monotone or not missing at random. Statistical methods in medical research. 2018;27:352–63. 10.1177/0962280216628902

21. Seaman SR, White IR. Review of inverse probability weighting for dealing with missing data. Statistical methods in medical research. 2013;22:278–95. 10.1177/0962280210395740

22. R Core Team. R: A language and environment for statistical computing. Vienna, Austria: R Foundation for Statistical Computing; 2024.

23. Elm EV, Altman DG, Egger M, Pocock SJ, Gøtzsche PC, Vandenbroucke JP. Strengthening the reporting of observational studies in epidemiology (STROBE) statement: Guidelines for reporting observational studies. Bmj. 2007;335:806–8. 10.1136/bmj.39335.541782.AD

24. Encalada S, Atchison JW, Prideaux CC, Narouze S, Mosquera-Moscoso J, De Mendonca LFP, et al. The association between chronic pain and metabolic syndrome: A scoping review. Pm&r. 2025;17:1107–19. 10.1002/pmrj.13361

25. Zhou WBS, Meng J, Zhang J. Does Low Grade Systemic Inflammation Have a Role in Chronic Pain? Frontiers in molecular neuroscience. 2021;14:785214. 10.3389/fnmol.2021.785214

26. Yoshimoto T, Ochiai H, Shirasawa T, Nagahama S, Uehara A, Sai S, et al. Sex differences in the association of metabolic syndrome with low back pain among middle-aged Japanese adults: A large-scale cross-sectional study. Biology of sex differences. 2019;10:33. 10.1186/s13293-019-0249-3

27. Sanchez-Santos M, Judge A, Gulati M, Spector T, Hart D, Newton J, et al. Association of metabolic syndrome with knee and hand osteoarthritis: A community-based study of women. Seminars in arthritis and rheumatism. 2019;48:791–8. 10.1016/j.semarthrit.2018.07.007

28. Park IY, Cho NH, Lim SH, Kim HA. Gender-specific associations between fat mass, metabolic syndrome and musculoskeletal pain in community residents: A three-year longitudinal study. Böttcher Y, editor. Plos one. 2018;13:e0200138. 10.1371/journal.pone.0200138

29. Mogil JS. Sex differences in pain and pain inhibition: Multiple explanations of a controversial phenomenon. Nature reviews neuroscience. 2012;13:859–66. 10.1038/nrn3360

30. Segal NA, Nilges JM, Oo WM. Sex differences in osteoarthritis prevalence, pain perception, physical function and therapeutics. Osteoarthritis and cartilage. 2024;32:1045–53. 10.1016/j.joca.2024.04.002

31. Sorge RE, Totsch SK. Sex Differences in Pain. Journal of neuroscience research. 2017;95:1271–81. 10.1002/jnr.23841

32. Fillingim RB, King CD, Ribeiro-Dasilva MC, Rahim-Williams B, Riley JL. Sex, Gender, and Pain: A Review of Recent Clinical and Experimental Findings. The journal of pain. 2009;10:447–85. 10.1016/j.jpain.2008.12.001

