## Supplementary Tables S1-S6 for "Sex-specific associations between metabolic dysregulation and knee pain: a 9-year population-based cohort study"

**Supplementary Table S1. Longitudinal associations between baseline cMetS and knee pain at 9-year follow-up: full set of crude and adjusted logistic regression models with and without cMetS-by-sex interaction**

| **Model** | **Adjustment** | **Effect** | **OR (95% CI)** | **P-value** |
|---|---|---|---|---|
| Model 1: no interaction | Crude | cMetS | 1.08 (0.98 to 1.18) | 0.106 |
| Model 1: no interaction | Adjusted | cMetS | 1.08 (0.98 to 1.18) | 0.110 |
| Model 2: interaction | Crude | cMetS (males) | 1.40 (1.17 to 1.68) | <0.001 |
| Model 2: interaction | Crude | cMetS (females) | 0.94 (0.83 to 1.06) | 0.336 |
| Model 2: interaction | Crude | cMetS × sex | 0.67 (0.54 to 0.83) | <0.001 |
| Model 2: interaction | Adjusted | cMetS (males) | 1.41 (1.17 to 1.69) | <0.001 |
| Model 2: interaction | Adjusted | cMetS (females) | 0.94 (0.84 to 1.07) | 0.356 |
| Model 2: interaction | Adjusted | cMetS × sex | 0.67 (0.54 to 0.84) | <0.001 |

Model 1: without interaction. Model 2: with a cMetS-by-sex interaction term.
Model 2 crude: Measure of interaction on multiplicative scale: ratio of ORs (95% CI) = 0.67 (0.54 to 0.83); P = <0.001
Model 2 adjusted: Measure of interaction on multiplicative scale: ratio of ORs (95% CI) = 0.67 (0.54 to 0.84); P = <0.001
Note: Crude models are unadjusted. Adjusted models include age and sex where appropriate. For interaction models, sex-specific effects are shown for males and females together with the multiplicative interaction term. Abbreviations: cMetS = continuous metabolic syndrome score; OR = odds ratio; CI = confidence interval.

**Supplementary Table S2. Sex-specific longitudinal associations of individual baseline components of cMetS with knee pain at 9-year follow-up**

| Exposure | N | Male OR (95% CI) | Male P-value | Female OR (95% CI) | Female P-value | Interaction ratio of ORs (95% CI) | Interaction P-value |
|---|---|---|---|---|---|---|---|
| Waist circumference (baseline residual z) | 248 | 3.37 (1.70 to 6.68) | <0.001 | 1.07 (0.73 to 1.59) | 0.716 | 0.32 (0.15 to 0.70) | 0.004 |
| Triglycerides (baseline residual z) | 245 | 1.46 (1.00 to 2.15) | 0.053 | 1.09 (0.83 to 1.43) | 0.546 | 0.74 (0.46 to 1.19) | 0.220 |
| HDL-C (baseline residual z) | 245 | 0.58 (0.35 to 0.97) | 0.038 | 1.30 (0.91 to 1.86) | 0.147 | 2.23 (1.20 to 4.17) | 0.011 |
| Glucose (baseline residual z) | 247 | 2.08 (1.01 to 4.27) | 0.047 | 0.85 (0.52 to 1.39) | 0.526 | 0.41 (0.17 to 0.98) | 0.044 |
| Systolic BP (baseline residual z) | 248 | 2.02 (1.22 to 3.35) | 0.007 | 0.57 (0.36 to 0.91) | 0.018 | 0.28 (0.14 to 0.56) | <0.001 |

Adjusted for: age; sex-specific estimates derived from models including a component-by-sex interaction term

Note: Odds ratios are reported per 1 SD increase in each baseline residualized metabolic component. The interaction term is presented as the ratio of odds ratios on the multiplicative scale. Abbreviations: OR = odds ratio; CI = confidence interval; HDL-C = high-density lipoprotein cholesterol; BP = blood pressure.

**Supplementary Table S3. Baseline characteristics of participants according to follow-up assessment status**

| Variable | Completed follow-up | Not assessed at follow-up | SMD |
|---|---|---|---|
| Participants, n | 250 | 1262 | |
| Age, years | 37.3 ± 5.9 | 36.5 ± 6.0 | 0.127 |
| Missing | 1 (0.4%) | 1 (0.1%) | |
| Female, n (%) | 129 (51.8%) | 710 (56.3%) | 0.090 |
| Missing | 1 (0.4%) | 1 (0.1%) | |
| BMI, kg/m² | 26.1 ± 4.7 | 26.1 ± 5.6 | 0.002 |
| Missing | 1 (0.4%) | 3 (0.2%) | |
| Waist circumference, cm | 86.7 ± 12.8 | 86.3 ± 14.3 | 0.029 |
| Missing | 1 (0.4%) | 4 (0.3%) | |
| Triglycerides, mmol/L | 1.26 ± 0.98 | 1.16 ± 0.82 | 0.111 |
| Missing | 3 (1.2%) | 49 (3.9%) | |
| HDL-C, mmol/L | 1.33 ± 0.32 | 1.33 ± 0.32 | 0.016 |
| Missing | 3 (1.2%) | 49 (3.9%) | |
| Glucose, mmol/L | 5.55 ± 0.57 | 5.60 ± 0.87 | 0.066 |
| Missing | 1 (0.4%) | 9 (0.7%) | |
| Systolic BP, mmHg | 119.9 ± 14.1 | 120.6 ± 15.7 | 0.045 |
| Missing | 1 (0.4%) | 7 (0.6%) | |
| MetS by IDF, n (%) | 70 (28.0%) | 304 (24.1%) | 0.089 |
| Missing | 0 (0.0%) | 0 (0.0%) | |
| cMetS | -0.16 ± 3.03 | -0.02 ± 3.07 | 0.045 |
| Missing | 4 (1.6%) | 57 (4.5%) | |
| Education level, n (%) | | | 0.132 |
| incomplete secondary | 4 (1.6%) | 22 (1.8%) | |
| secondary or vocational | 69 (27.7%) | 464 (37.0%) | |
| higher education | 176 (70.7%) | 769 (61.3%) | |
| Missing | 1 (0.4%) | 7 (0.6%) | |
| Employment status, n (%) | | | 0.073 |
| employed | 214 (85.9%) | 1001 (79.9%) | |
| student | 0 (0.0%) | 4 (0.3%) | |
| homemaker | 23 (9.2%) | 159 (12.7%) | |
| unemployed | 10 (4.0%) | 78 (6.2%) | |
| retired/disabled | 2 (0.8%) | 11 (0.9%) | |
| Missing | 1 (0.4%) | 9 (0.7%) | |
| Self-rated health, n (%) | | | 0.053 |
| very good | 8 (3.2%) | 26 (2.1%) | |
| good | 96 (38.6%) | 439 (35.0%) | |
| fair | 131 (52.6%) | 684 (54.5%) | |
| poor | 12 (4.8%) | 100 (8.0%) | |
| very poor | 2 (0.8%) | 5 (0.4%) | |
| Missing | 1 (0.4%) | 8 (0.6%) | |
| Physical activity, n (%) | | | 0.108 |
| yes | 98 (39.4%) | 388 (30.9%) | |
| sometimes | 36 (14.5%) | 206 (16.4%) | |
| no | 115 (46.2%) | 661 (52.7%) | |

| | | | |
|---|---|---|---|
| Missing | 1 (0.4%) | 7 (0.6%) | |
| Smoking status, n (%) | | | 0.109 |
| never smoked | 100 (40.2%) | 487 (38.8%) | |
| former smoker | 82 (32.9%) | 326 (26.0%) | |
| current smoker | 67 (26.9%) | 442 (35.2%) | |
| Missing | 1 (0.4%) | 7 (0.6%) | |

Note: Continuous variables are presented as mean ± standard deviation and categorical variables as number (%). Missing values are shown for all variables. Standardized mean differences (SMDs) were used to compare baseline characteristics between participants who completed the follow-up assessment and those not assessed at follow-up. This table is intended to evaluate potential selection bias. MetS by IDF was defined using clinical and laboratory criteria; information on antihypertensive treatment was not included. Abbreviations: BMI = body mass index; HDL-C = high-density lipoprotein cholesterol; BP = blood pressure; MetS = metabolic syndrome; cMetS = continuous metabolic syndrome risk score; IDF = International Diabetes Federation; SMD = standardized mean difference.

**Supplementary Table S4. Predictors of follow-up participation and inverse probability weights**

| Variable | Primary model OR (95% CI) | Primary model P-value | Additional model OR (95% CI) | Additional model P-value |
|---|---|---|---|---|
| Intercept | 0.21 (0.03 to 1.64) | 0.132 | 0.36 (0.05 to 2.56) | 0.298 |
| Age (per year) | 1.03 (1.01 to 1.06) | 0.009 | 1.03 (1.00 to 1.05) | 0.041 |
| Female (vs male) | 0.85 (0.59 to 1.20) | 0.353 | 0.77 (0.55 to 1.06) | 0.113 |
| Waist circumference, cm | 1.01 (0.99 to 1.02) | 0.405 | 1.00 (0.99 to 1.02) | 0.657 |
| Triglycerides, mg/dL | 1.00 (1.00 to 1.00) | 0.056 | 1.00 (1.00 to 1.00) | 0.064 |
| HDL-C, mg/dL | 1.00 (0.99 to 1.02) | 0.510 | 1.00 (0.99 to 1.02) | 0.453 |
| Glucose, mmol/L | 0.85 (0.66 to 1.05) | 0.178 | 0.85 (0.67 to 1.04) | 0.162 |
| Systolic BP, mmHg | 0.99 (0.98 to 1.00) | 0.104 | 0.99 (0.98 to 1.00) | 0.083 |
| Higher education (vs no higher education) | 1.48 (1.08 to 2.04) | 0.015 | | |
| Not employed/other (vs employed) | 0.72 (0.48 to 1.05) | 0.100 | | |
| Self-rated health: fair (vs good/very good) | 0.89 (0.66 to 1.20) | 0.443 | | |
| Self-rated health: poor/very poor (vs good/very good) | 0.68 (0.35 to 1.21) | 0.210 | | |
| Physical activity: sometimes/no (vs yes) | 0.73 (0.54 to 0.98) | 0.036 | | |
| Ever smoker (vs never) | 1.04 (0.77 to 1.41) | 0.794 | | |

| | | | |
|---|---|---|---|
| N in response model | 1446 | | 1451 |
| Responders | 246 | | 246 |
| Non-responders | 1200 | | 1205 |
| IPW: min | 2.29 | | 2.03 |
| IPW: 25th percentile | 4.90 | | 5.30 |
| IPW: median | 6.10 | | 5.99 |

| | | | |
|---|---|---|---|
| IPW: mean | 6.66 | | 6.17 |
| IPW: 75th percentile | 7.64 | | 6.85 |
| IPW: max | 45.56 | | 37.12 |

Note: Odds ratios (ORs) are derived from logistic regression models of follow-up participation.
The primary response model included age, sex, waist circumference, triglycerides, HDL-C, glucose, systolic blood pressure, as well as education, employment status, self-rated health, physical activity, and smoking (recoded categories).
A simpler cardiometabolic model including age, sex, waist circumference, triglycerides, HDL-C, glucose, and systolic blood pressure was used as an additional sensitivity analysis.
Inverse probability weights (IPW) were calculated as the inverse of the predicted probability of follow-up participation.

**Supplementary Table S5. Sensitivity analyses of the association between baseline cMetS and knee pain using inverse probability weighting**

| Analysis | N | Knee pain / no knee pain by sex, n | Male OR (95% CI) | Male P-value | Female OR (95% CI) | Female P-value | Interaction ratio of ORs (95% CI) | Interaction P-value |
|---|---|---|---|---|---|---|---|---|
| Main analysis (unweighted) | 245 | Male: 26/93; Female: 45/81 | 1.41 (1.17 to 1.69) | <0.001 | 0.94 (0.84 to 1.07) | 0.356 | 0.67 (0.54 to 0.84) | <0.001 |
| IPW-weighted analysis, extended socio-clinical response model | 245 | Male: 26/93; Female: 45/81 | 1.35 (1.25 to 1.45) | <0.001 | 0.94 (0.89 to 0.99) | 0.017 | 0.70 (0.64 to 0.76) | <0.001 |
| IPW-weighted analysis, cardiometabolic response model | 245 | Male: 26/93; Female: 45/81 | 1.43 (1.33 to 1.55) | <0.001 | 0.92 (0.87 to 0.97) | 0.002 | 0.64 (0.58 to 0.70) | <0.001 |

Note: Odds ratios represent the association between baseline cMetS and knee pain per 1-unit increase in cMetS. Sex-specific estimates were derived from logistic regression models including a cMetS × sex interaction term and adjusted for age. IPW analyses were performed as sensitivity analyses using two alternative response models: a socio-clinical response model and a simpler cardiometabolic response model. The interaction term is presented as the ratio of odds ratios on the multiplicative scale.

**Supplementary Table S6. BMI-adjusted sensitivity analysis**

| Sex | Knee pain, n | No knee pain, n | OR per 1-unit increase in cMetS (95% CI) | P-value |
|---|---|---|---|---|
| Male | 26 | 93 | 1.34 (1.10 to 1.64) | 0.003 |
| Female | 45 | 81 | 0.90 (0.78 to 1.05) | 0.171 |

Measure of interaction on multiplicative scale: ratio of ORs (95% CI) = 0.67 (0.54 to 0.84); P = <0.001

Adjusted for: age, BMI

Note: Odds ratios represent the longitudinal association between baseline cMetS and knee pain at 9-year follow-up per 1-unit increase in cMetS. Abbreviations: cMetS = continuous metabolic syndrome score; OR = odds ratio; CI = confidence interval; BMI = body mass index.
